# Validity and Reliability of a Behavior Assessment Questionnaire for Children with Obesity

**DOI:** 10.1101/2021.05.12.21256827

**Authors:** So Yeon Paek, Lonnie C. Roy, Mark J. DeHaven, Elyse Carson, Sarah E. Barlow, LeAnn Kridelbaugh

## Abstract

The 10-item Behavior Assessment Questionnaire (BAQ) was developed to assess parent-report of child screen time, physical activity, and food consumption during the past 3 months in children with obesity. Response options were on a 5-point scale, converted to 0-100, with higher scores indicating healthier behavior. To evaluate, two convenience samples of parents of children 5-18 years completed the questionnaire: a cohort presenting to an obesity program (n=83) and a cohort of community events attenders and hospital employee parents (n=147). Scores had a normal distribution without floor or ceiling effects. Cronbach’s alpha for the 10-item scale was .71. Factor analysis yielded three component factors with Cronbach’s alpha of .66, .75, and .59 for the Screen Time, Physical Activity, and Food Consumption dimensions respectively. Scores of the obesity group (49.02 [SD 14.52]) were lower than scores of the community group (55.44 [SD 13.55]), p=.001. The BAQ demonstrated reliability and validity for use as an index of lifestyle behaviors.

**Key Messages:**

1. A short, easily administered questionnaire, completed by parents, about obesity-associated lifestyle behaviors of children demonstrates reliability and validity
2. This questionnaire could help pediatric providers assess behaviors in order to provide more targeted lifestyle counseling for children to reduce obesity risk.

## Introduction

Pediatric obesity is epidemic in the United States and throughout the world. The most current U.S. prevalence data indicate that over 19% of children and adolescents have obesity, and over 5% have severe obesity.^1^ Many of these children suffer from co-morbid health complications, such as type 2 diabetes and metabolic syndrome, or are at risk for developing such health conditions later in life.^2^

Research has demonstrated that a multidisciplinary family-based approach is effective in treating pediatric overweight and obesity.^3^ Effective comprehensive programs address physical activity, healthy eating and behavior change. A twelve-week program at Children’s Medical Center in Dallas, Texas, was developed based on these principles and a literature review.^4-9^ The curriculum focused on reduced television watching, increased physical activity, and consumption of a healthy, well-balanced diet. Program participants were children between the ages of 6-18 years with a BMI greater than or equal to the 95^th^ percentile for age, at least one family member committed to attend class and participate in the program with the child.

Despite evidence supporting the benefits of regular physical activity and a healthy diet with limited energy dense foods and beverages, availability of instruments to measure healthy lifestyle in children is very limited. The measures registry from the National Center for Childhood Obesity Research.^10^ whose members include the Centers for Disease Control and Prevention and the National Institutes of Health, reveals a gap: limitations of registry measures include length,^11,12^ focus on parenting strategies and self-efficacy rather than child behavior,^13,14^ or are not self-administered.^15^ One questionnaire was short and self-administered but evaluated on a small sample of 35 general pediatric patients.^16^

To address the need for a behavior assessment questionnaire to supplement the anthropometric measure, the 10-item Behavior Assessment Questionnaire (BAQ) was developed. The present study evaluates the effectiveness of the BAQ in determining lifestyle behaviors as indicators of obesity. We report the validity and reliability of the instrument in supporting its utility as a measure of eating behavior and activity levels.

## Methods

### Questionnaire

Question items were developed following a comprehensive literature review and comprise 3 items that address television watching as a component of pediatric obesity management; 2 items on physical activity; and 5 items on food consumption and eating habits that affect weight. The questionnaire asks for parent’s perceptions of their child’s habits and activities in the past 3 months. The answers range from 0 to 4 in a 5-point response scale, with higher scores denoting more desirable behaviors.

### Participants

Convenience samples of two groups of English-speaking parents of children ages 5 to 18 years were recruited. Group 1 were parents of children undergoing initial assessment of obesity at the medical center following referral by their primary care physician. Group 2 were participants at local community health fairs and Children’s Medical Center employees. Data were gathered using a self-administered paper survey. Parents also reported child’s age and sex. BMI z score was calculated in group 1 using the CDC formula, based on measured weights and heights. Weights and heights of children in group 2 were not obtained; community prevalence of healthy weight, overweight, and obesity was presumed.

### Statistics

Feasibility was examined by evaluating the percentage of missing data for each item and percentage of the missing values in the overall questionnaire. Overall scores were computed as the average of the items answered. The range of measurement and variability of the instrument were evaluated by calculating the percentage of responses for each item at the minimum (percentage floor) and maximum (percentage ceiling) response category, as well as item means and standard deviations. Response frequencies (number of times specific responses were selected) were examined through measurements of central tendency (mean) and variability (standard deviation). Questionnaires with complete data were used for reliability and validity testing. Cronbach’s coefficient “alpha” was calculated to assess internal consistency. Construct validity was evaluated by factor analysis and known-groups method (independent sample t-test). Items were weighted to 100pt scale for ease of interpretation (0=0, 1=25, 2=50, 3=75, 4=100). The questionnaire is shown in Figure 1.

**Figure 1:**
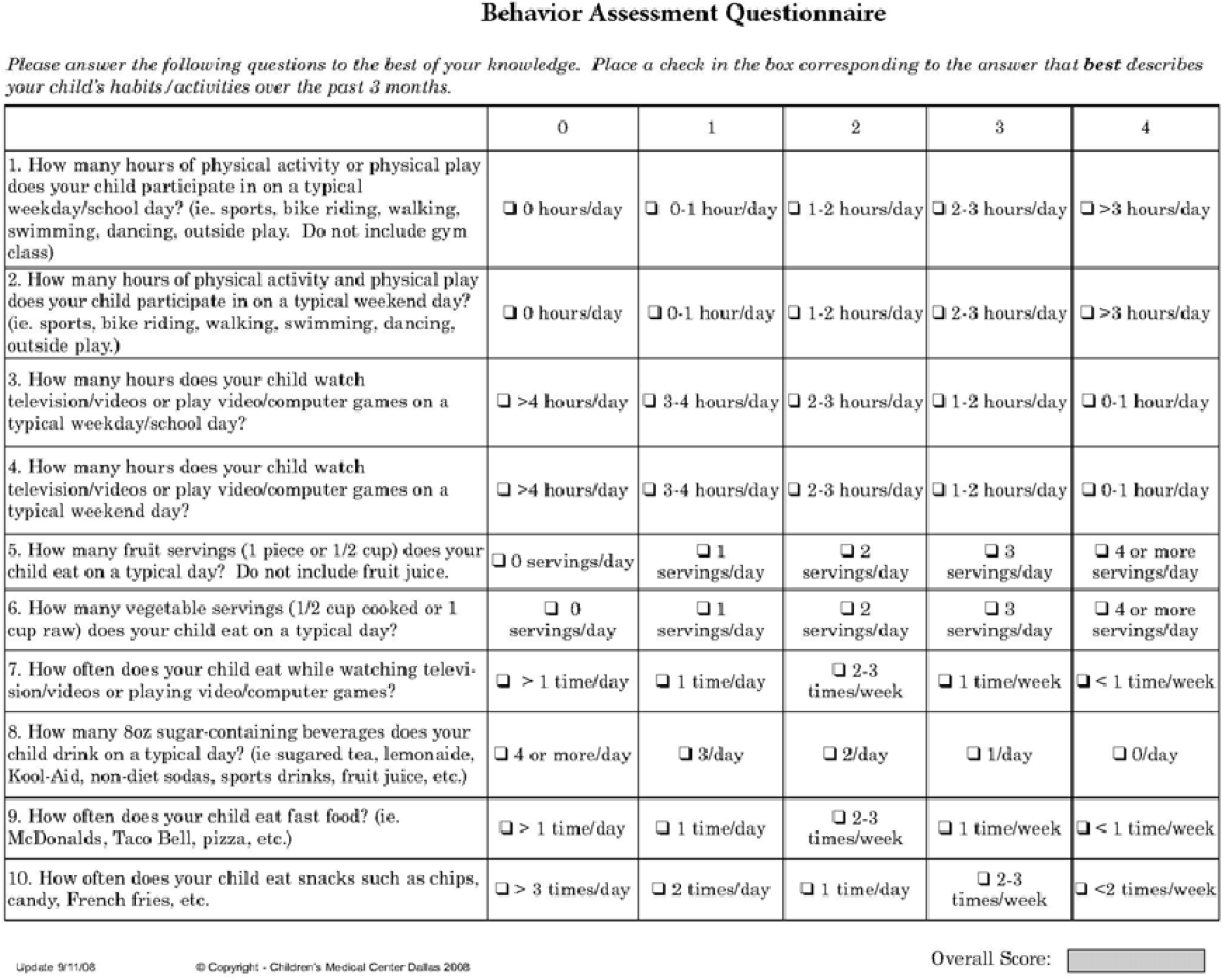
Behavior Assessment Questionnaire

## Results

Questionnaires were completed by 83 parents of children undergoing obesity evaluation (group 1) and 147 parents from the community (group 2). The average age of the 104 males (45%) and 126 females (55%) was 10.40 years (SD = 3.35) with a range of 5 to 18 years. The children undergoing obesity evaluation (group 1) were younger (mean age 9.2 vs. 11.8 years; p< .001) and more likely to be female (63% vs. 50%; p = .072) than the community sample. Of the 83 children in group 1, 75 had calculable BMI z-scores. Z-scores ranged from 1.71 to 3.38, with mean of 2.49, median of 2.54, and SD = 0.35.

The percentage of missing values in our analysis ranged from 0.4% (Q5, Q10) to 2.2% (Q6, Q7), with an overall questionnaire missing of 1.1%. Responses were observed for all response categories of each questionnaire item. Floor and ceiling effects were not substantial in our study. However, there was a tendency for Q3, Q8, Q9, and Q10 to be positively skewed and Q4 to be negatively skewed. Table 1 presents floor, ceiling, and missing frequency.

**Table 1:**
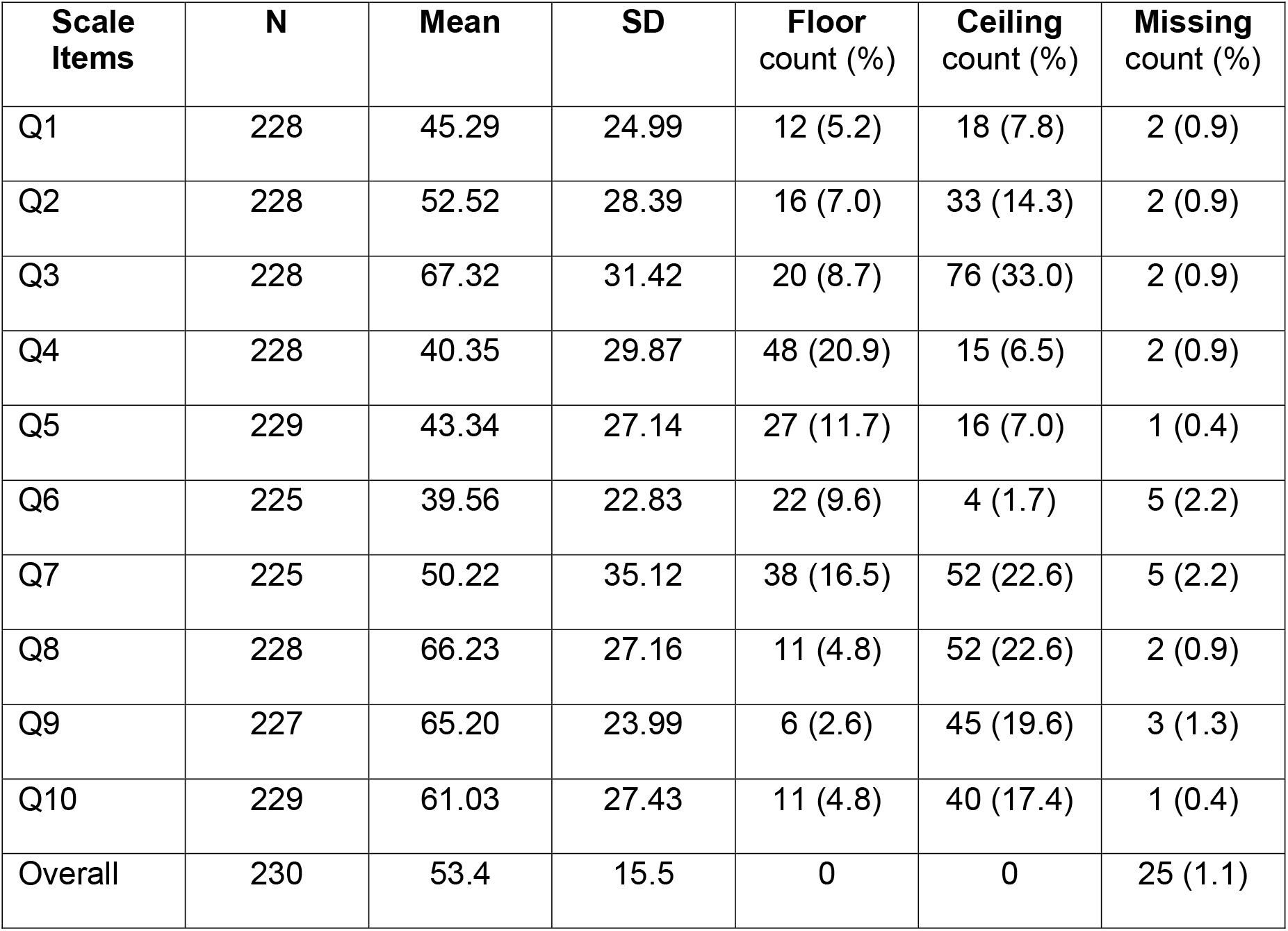
Frequency of floor, ceiling, and missing responses for Behavior Assessment Questionnaire (BAQ). Floor and ceiling refer to the minimum (0) and maximum (100) response categories to an item or overall score, on a 100-point scale.

| **Scale Items** | **N** | **Mean** | **SD** | **Floor count (%)** | **Ceiling count (%)** | **Missing count (%)** |
| --- | --- | --- | --- | --- | --- | --- |
| Q1 | 228 | 45.29 | 24.99 | 12 (5.2) | 18 (7.8) | 2 (0.9) |
| Q2 | 228 | 52.52 | 28.39 | 16 (7.0) | 33 (14.3) | 2 (0.9) |
| Q3 | 228 | 67.32 | 31.42 | 20 (8.7) | 76 (33.0) | 2 (0.9) |
| Q4 | 228 | 40.35 | 29.87 | 48 (20.9) | 15 (6.5) | 2 (0.9) |
| Q5 | 229 | 43.34 | 27.14 | 27 (11.7) | 16 (7.0) | 1 (0.4) |
| Q6 | 225 | 39.56 | 22.83 | 22 (9.6) | 4 (1.7) | 5 (2.2) |
| Q7 | 225 | 50.22 | 35.12 | 38 (16.5) | 52 (22.6) | 5 (2.2) |
| Q8 | 228 | 66.23 | 27.16 | 11 (4.8) | 52 (22.6) | 2 (0.9) |
| Q9 | 227 | 65.20 | 23.99 | 6 (2.6) | 45 (19.6) | 3 (1.3) |
| Q10 | 229 | 61.03 | 27.43 | 11 (4.8) | 40 (17.4) | 1 (0.4) |
| Overall | 230 | 53.4 | 15.5 | 0 | 0 | 25 (1.1) |

The mean overall score was 53.4 with a standard deviation of 15.5. Distribution of the BAQ overall scale scores approached normality, yielding an adequate level of variance in scores, as shown in Figure 2. No floor or ceiling effects were observed in the overall scale score.

**Figure 2.**
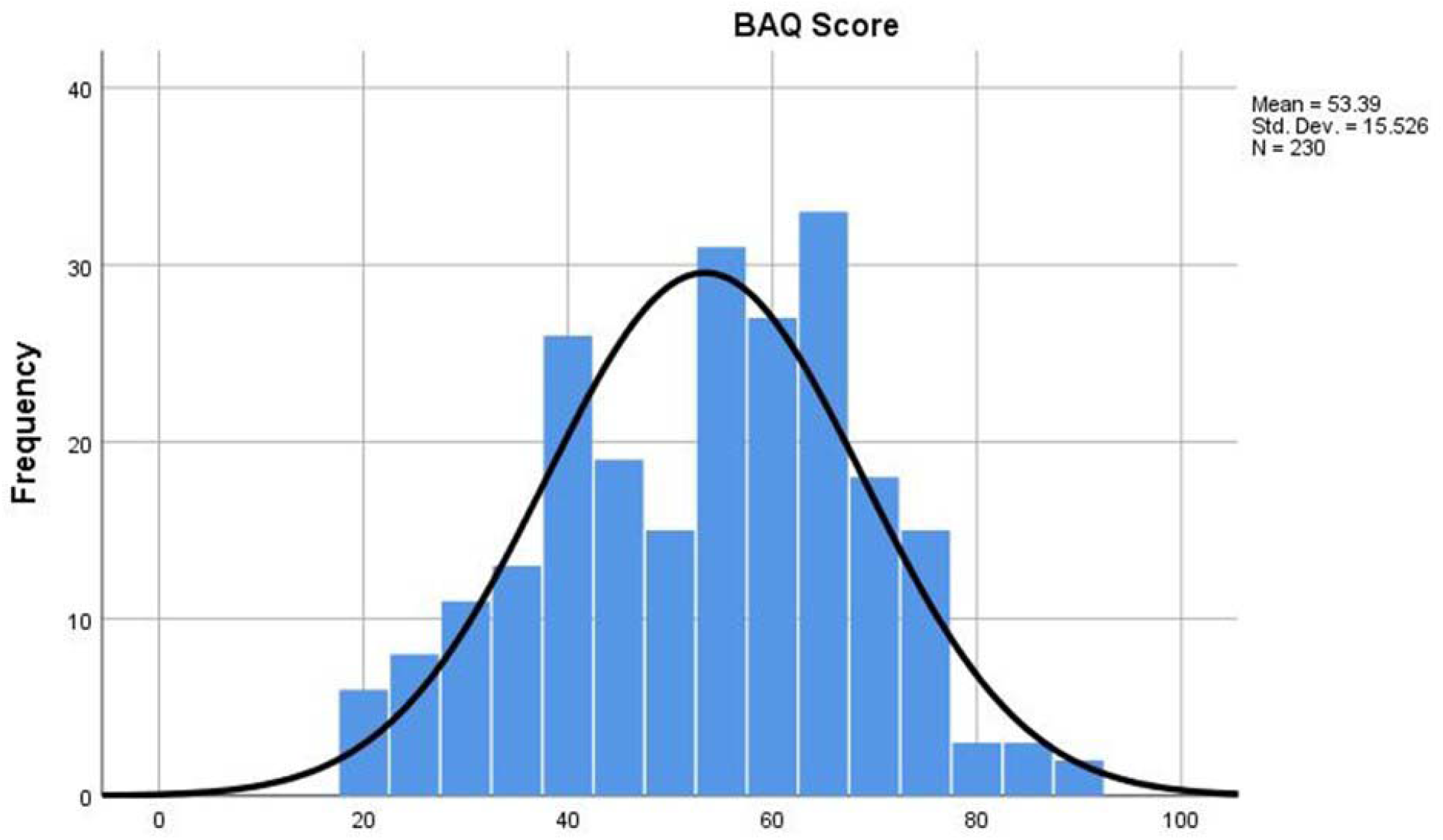
Distribution of the Behavior Assessment Questionnaire (BAQ) overall scale scores. The distribution approaches normality, yielding an adequate level of variance. No floor or ceiling effects were observed in the overall scale score.

Factor analysis of the questionnaire yielded a three-component factor structure as originally hypothesized, although the loading of the “sugar drinks” item on the Screen Time dimension was not anticipated (Table 2). The three factors explained 57% of the overall variance of the items. All items loaded above 0.60 in the Screen Time dimension and above 0.80 in the Physical Activity dimension, suggesting a high degree of inter-correlation of the items with the respective dimension. The Food Consumption dimension generated four well inter-correlated items, although not as strongly inter-correlated as the other two factors. Table 2 also displays measures of item redundancy. Multiple factor loadings above 0.90 in a dimension suggest that two questions are redundant. No items in our questionnaire indicated redundancy within dimensions.

**Table 2.** Factor loadings and Cronbach’s reliability coefficients for Behavior Assessment Questionnaire (BAQ)

| **n=212** | **Overall** | **Screen Time** | **Physical Activity** | **Food Consumption** |
| --- | --- | --- | --- | --- |
| Q4 Hours Watch TV Weekend |  | 0.734 |  |  |
| Q3 Hours Watch TV Weekday |  | 0.710 |  |  |
| Q7 Frequency Eat While Watching TV |  | 0.647 |  |  |
| Q8 Sugar Drinks Daily |  | 0.611 |  |  |
| Q2 Hours per Weekend Physical Activity |  |  | 0.851 |  |
| Q1 Hours per Weekday Physical Activity |  |  | 0.838 |  |
| Q5 Fruit Servings Daily |  |  |  | 0.821 |
| Q6 Vegetable Servings Daily |  |  |  | 0.684 |
| Q10 Frequency Eat Unhealthy Snacks |  |  |  | 0.538 |
| Q9 Frequency Eat Fast Foods |  |  |  | 0.497 |
| CRONBACH'S ALPHA | 0.711 | 0.657 | 0.749 | 0.594 |

\*NOTE: Factor loadings less than 0.40 are suppressed in the table

Reliability and validity testing were performed on the 212 (92%) complete questionnaires. The reliability for all questions (α = 0.711, Table 2) exceeded 0.70, the standard value for good reliability. Reliability among scale sub-dimensions varied. Screen Time (α = 0.657) and Physical Activity (α = 0.749) components potentially support further sub-dimension analysis, while Food Consumption (α = 0.594) did not meet acceptable reliability for sub-dimension analysis. The overall BAQ self-report scores of the community sample demonstrated a higher (better) mean score than the obesity evaluation clinic sample (55.44 vs. 49.02, p=.003), evidence of divergent validity. No significant sex effect was evident when comparing the two groups (data not shown).

## Discussion

The Behavior Assessment Questionnaire (BAQ) was developed using existing literature, for the purpose of measuring lifestyle behaviors among children with obesity. The small values for percentage missing support the feasibility of the self-administered format. The overall questionnaire score yielded good reliability for using the instrument as an index of lifestyle behaviors, and it discriminated between known groups of children with obesity and a child population sample. Sub-scale analysis for the Screen Time and Physical Activity components was marginally supported, and although the Food Consumption dimension did not meet the minimum acceptable reliability values for sub-scale use, it may be of use for further exploratory analysis. The unexpected loading of the “sugar drinks” item in the Screen Time dimension of the BAQ may reflect the association of higher screen time with increased calories.^17^ Further research is needed to determine the utility of the sub-scales as independent measures.

As predicted, the overall BAQ score was significantly different between a group of children with known high BMI and children of the general population. Known-groups analysis yielded results with a lower mean value for the obesity clinic sample participants than the population sub-sample, suggesting that the children with obesity demonstrated behaviors that put them at greater risk for obesity. Further, the behaviors represented in the instrument are those recommended as targets for intervention with children with obesity.^6^ Thus, the instrument has potential utility as a pre-and post-intervention measure of behavior change in obese children.

Limitations of this study include its small sample size. Additionally, the weight of the children in the community sample collected at health fairs and among children of hospital employees was unknown. Some differences in age and sex in the two groups may have affected results.

Despite these limitations, our findings indicate that the BAQ is feasible, and the good reliability and validity for the summary score make it a useful instrument. A variety of community and clinical programs targeting families with children with obesity could find this measure an easily administered, useful assessment of eating and activity behaviors.

## Data Availability

The data are not available.

